# Mathematical Modeling of COVID-19 Pandemic in the African Continent

**DOI:** 10.1101/2020.10.10.20210427

**Authors:** Nawel Aries, Houdayfa Ounis

## Abstract

The present work aims to give a contribution to the understanding of the highly infectious pandemic caused by the COVID-19 in the African continent. The study focuses on the modelling and the forecasting of COVID-19 spread in the most affected African continent, namely: Morocco, Algeria, Tunisia, Egypt and South Africa and for the sake of comparison two of the most affected European country are also considered, namely: France and Italy. To this end, an epidemiological SEIQRDP model is presented, which is an adaptation of the classic SIR model widely used in mathematical epidemiology. In order to better coincide with the preventive measures taken by the governments to deal with the spread of COVID-19, this model considers the quarantine. For the identification of the model’s parameters, official data of the pandemic up to August 1^st^, 2020 are considered. The results show that the number of infections due to the use of quarantine is expected to be very low provided the isolation is effective. However, it is increasing in some countries with the early lifting of containment. Finally, the information provided by the SEIQRDP model could help to establish a realistic assessment of the short-term pandemic situation. Moreover, this will help maintain the most appropriate and necessary public health measures after the lockdown lifting.

## 1. Introduction

The year 2020 is and will be forever marked by the COVID-19 pandemic caused by the highly infectious disease SARS-CoV-2. According to the World Health Organization, since its first appearance in late 2019 in the Wuhan region of China, the Covid-19 has affected more than 6 million people and has caused the death of almost 400 thousand people worldwide, during the first half of 2020. However, the spread of the disease and the damage caused by it did not happen in the same way and at the same time for all continents. Indeed, taking the case of the African continent, the COVID-19 pandemic reached a significant point by exceeding 180,000 cases in early June with more than 5,000 deaths. Although the pandemic has reached a worrying threshold, the spread of the disease in Africa has not followed the exponential path as in the rest of the world (e.g. Europe and United States). According to WHO’s analysis, the relatively low mortality rate compared to other continents is likely due to the demographic nature of the continent. Indeed, demographically, Africa is considered as the youngest continent, with more than 60% of the population under the age of 25. However, the disease still represents a danger for the African population, particularly, the South and African North. In fact, according to the WHO, the above-mentioned regions account for more than 56% of the cases recorded in Africa.

To date, in the absence of an effective vaccine or treatment against COVID-19, African governments have taken several measures to limit the spread of the disease as much as possible (e.g. quarantine, social distancing, lockdown, curfew, masks, etc.). Furthermore, during this anti-COVID-19 battle, in addition to medical and biological research, theoretical studies based on mathematical models can also play an important role in understanding the pandemic dynamics (e.g. predicting the inflection point and of the end time). These models are therefore an important decision-making tool for coping with this pandemic. Since the apparition of the disease, the literature has witnessed an avalanche of work on modelling and predicting the behaviour of the COVID-19 pandemic. However, most of these works have focused on China [1-5], Europe [6-11] and the United States [7, 11-13], which is understandable because these are the regions which were the first and most affected by COVID-19. Nevertheless, studies on the African continent are practically negligible compared to those dedicated to the above-mentioned regions [14-16].

The present paper aims to present a contribution on the modelling and the forecasting of COVID-19 spread in the most affected African continent, namely: Morocco, Algeria, Tunisia, Egypt and South Africa. In addition, for the sake of comparison two European countries are considered, namely: France and Italy. Thereby, an epidemic SEIQRDP model is presented, which is an adaptation of the classic SIR model widely used in mathematical epidemiology. In the next section, we present the data and the model. Section 3 presents the results of the application of the model for the above-mentioned countries through the estimation of the epidemic parameters and the adjustment of the model. The discussion of the results obtained as well as a forecast analysis, are presented in the same section. The last section provides some future directions for using the SEIQRDP model to control and prevent the spread of the COVID-19virus.

### 2. Data and Model

#### 2.1. Data

The epidemiologic data of Covid-19 were provided by the Johns Hopkins University Center for Systems Science and Engineering (JHUCSSE) [17], and derived from the official case count from the World Health Organization. The data provides the total number of confirmed, quarantined, recovered and death cases, for each country since January 22, the day that WHO announced an epidemic caused by a new coronavirus in China. Our preliminary study includes the most affected African countries and one of the main endemic foci in Europe, namely: Algeria, Morocco, Egypt, South Africa, Tunisia, France, and Italy. Figure 1 shows the cumulative reported cases of COVID-19 in the considered countries. The data refer to daily cumulative cases from January 22, 2020 until August 1st, 2020. It is important to note that these data are influenced by the capacity and strategy of countries in case detection. Some countries perform more tests than others. However, the provided data still provide indicator for tracking the trajectories of the epidemic. It can be well seen from figure 1 that the curve of newly detected cases in European countries is gradually flattening. On the other hand, there is an epidemic surge in the African countries, where the number of cumulative cases shows a different picture. Indeed, South Africa and Egypt are currently the countries with most known confirmed and deaths cases and the curve of infections still pointing mostly upwards.

**Figure 1.**
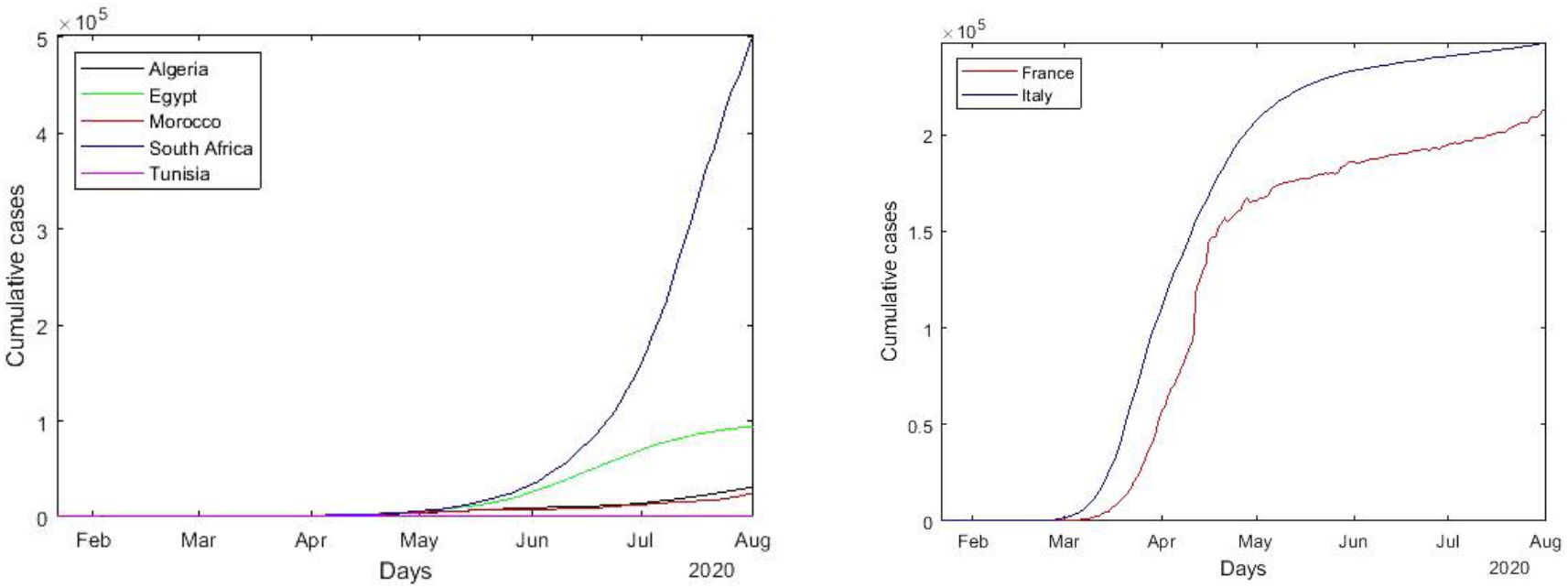
Cumulative reported cases of COVID-19 disease.

Unlike the last ones, the curve of cumulative cases is increasing less rapidly in Algeria and Morocco. Although Algeria has the highest number of deaths ahead the African countries. While, the curve of newly detected cases in Tunisia quickly flattened. Infection numbers have, however, also shown some signs of slowing down. It should be noted that the African continent is the last to be affected by the coronavirus. Indeed,

The epidemic spread to Africa a few weeks after Europe, allowing its leaders to adopt preventive measures well in advance. South Africa, Tunisia, Morocco and Algeria imposed the lockdown and curfews before the epidemic had had time to spread. In addition, the low population density has considerably limited the transmission of the virus. Indeed, the inhabitants are generally concentrated in the capitals.

#### 2.2. SEIQRDP Model

Using a pandemic modeling makes it possible to understand, describe and forecast its behavior and its spread. Indeed, the mathematical models assess hypotheses, indicate trends, and help develop public health responses by estimating risks, in real time, during an epidemic. Nevertheless, modeling is a simplified representation of reality. Its precision is limited here by the ignorance of certain factors and mechanism of propagation of COVID-19.

In this work, the SEIQRDP epidemiological model with seven different components, proposed by [2] is used. The model is an adaptation of the classic SEIR model [18, 19], widely used to study the COVID-19 pandemic in many countries with variations in components and parameters for adaptations to regions and study period [16, 19-23]. The present model considers the effect of quarantine adopted by many countries as an effective means of preventing the spread. The stochastic SEIQRDP model is specified by the following system of differential equations:

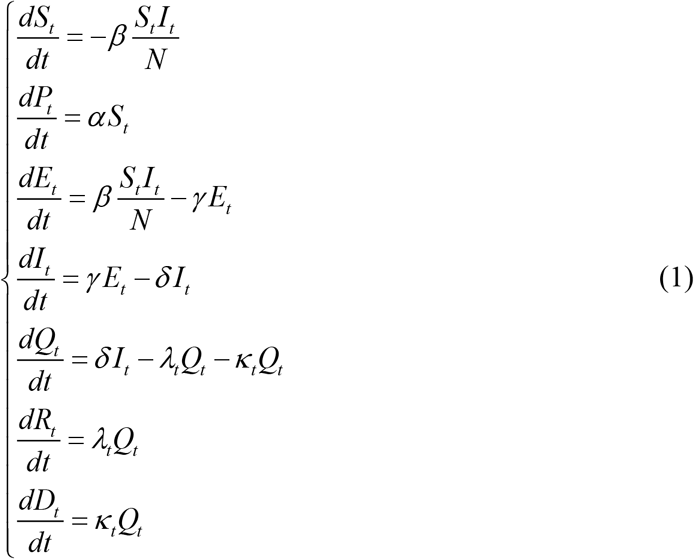

Where the different component at time *t*, i.e, *S*_*t*_, *P*_*t*_, *E*_*t*_, *I*_*t*_, *Q*_*t*_, *R*_*t*_, *D*_*t*_ are respectively, the number of susceptible cases (individuals who can contract the disease), the number of protected cases, who have become insensitive to the disease, mainly by following the recommended protective measures, the number of exposed case (individuals that have contracted the disease but not yet infectious), the number of infectious cases (infectious individuals but not yet quarantined), the number of quarantined cases, i.e active cases (confirmed and infected), the recovered and death cases (individuals removed from the chain of transmission), respectively. The component of protected part of people, namely, *P*_*t*_, was introduced to reflect the growing awareness of the population while respecting the lockdown. It should be noted that the total population N is assumed to be homogenous, i.e. there is no birth or death and recovered people remain immune once they recover from the disease where, *N* = *S*_*t*_ + *P*_*t*_ + *E*_*t*_ + *I*_*t*_ + *Q*_*t*_ + *R*_*t*_ + *D*_*t*_.

The protection rate *α*, represents the population that takes into consideration security measures and the actions of health authorities, it was introduced assuming that the sensitive population decreases steadily. Moreover, beside the parameter *α*, all the remaining parameters depend on the evolution of the epidemic, the health care and the screening capacities and are calculated based on the daily numbers of confirmed, recovery and death cases. The infection rate *β*, represent the average number of contacts per-capita per time, multiplied by the probability of successfully getting infected when coming into contact with an infected individual. While, the latent rate *γ*^−1^, represent the average time for a latent individual to become infectious, which is estimated within several days [4, 24]. Furthermore, the quarantined rate *δ*^−1^ represent the average time for an infectious individual to enter in quarantine, which is considered to be between 2 and 14 days [2]. Finally, to consider the measures taken by the different countries in this study, we assume that the recovery rate *λ*_*t*_ and the death rate *κ*_*t*_ are time dependent function, as confirmed by [2, 21].

In case of a new disease as the COVID-19, the medical staff must learn new therapeutic procedures and treat patients with new symptoms every day. Hence, the time of recovery, cannot be a constant, because the recovery time at the start of the disease is longer, this means a slower recovery rate which gradually increasing with time, considering the measures taken by the governments. Consequently, we will assume that the recovery rate is modeled by following function,

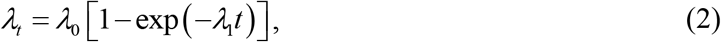

where *λ*_0_ and *λ*_1_ are the fitted coefficients [2].

Unlike the recovery rate, the mortality rate quickly decreases over time. This is due to the medical assistance, the adaptation of the pathogen and the development of new treatments. Hence, we will assume that the mortality rate is modeled by an exponential decay function given as follows,

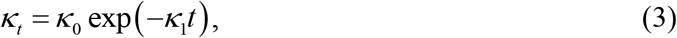

where *κ*_0_ and *κ*_1_ are the fitted coefficients [2].

However, one of the most discussed parameters in the current COVID-19 pandemic is the basic reproductive number [2, 24-26]. Biologically, this number, represented by *R*_0_, corresponds to the number of new infections caused by an infected individual in a susceptible population. One of the main reasons of the importance of the basic reproduction number is that it determines the fraction of the population that needs to be immunized for the epidemic to start to decline. The reproductive number will vary during an epidemic for two reasons. On the one hand, the public health measures are put in place, like the lockdowns, social distancing, and other mitigation strategies to keep the case count and death toll low in this pandemic. On the other hand, the epidemic spreads and people who have recovered are protected against reinfection (at least in the near future). Thus, the population becomes immune.

If the *R*_0_ is above 1, each infection breeds more, and the outbreak will continue to grow. When it falls below 1, the outbreak will continue but at a lower death rate.

In this study, the R_0_ is calculated by the next generation matrix method [22], based on a endemic equilibrium, given by : *S* = *N, P* = *E* = *I* = *Q* = *R* = *D* = 0. We must solve the equation, *R*_0_ = *ρ* (*FV*^−1^), where *F* and *V* are, respectively, the derivatives of the new infections matrix *ℱ* and the transition matrix *𝒱*, evaluated at the endemic equilibrium and is given by, *R*_*t*_ = *β * δ*^−1^(1 − *α*)^*t*^, where *t* represent the number of days.

The nonlinear system of the SEIQRDP model given by (1.1) can only be solved using numerical methods to observe the dynamics of the model. Firstly, we estimated the parameters, namely, *α, β, γ*^−1^,*δ*^−1^, *λ*_*t*_, *κ*_*t*_. In this step, we search the unknown parameters for the SEIQRDP model by global optimization. This is done automatically by implementing the nonlinear least square regression method. The optimization of the model parameters to describe the experimental results was performed via the minimization of the sum of the squared errors between the epidemiologic and the modeling data, during successive iterations, which finally gives the best parameters for data fitting. It should be noted that the use of the cumulative data should be avoided, since the cumulative incidence data are highly correlated, and the least squares approaches relies on the fitting of independent data. Therefore, the use of daily new data should be preferred, since these data are independent. Once all the parameters are known, we can solve the system of differential equations (1.1).

## 3. Results and Discussion

The estimated parameters for the considered countries are given in Table 1, where we consider the mean values of the recovery rate and deaths rate, *λ*_*t*_ and *κ*_*t*_, respectively.

**Table 1.** Parameter estimation of the SEIQRDP model

| Country | N | $\alpha$ | $\beta$ | $\gamma$ | $\delta$ | $\lambda_t$ | $\kappa_t$ |
| --- | --- | --- | --- | --- | --- | --- | --- |
| Morocco | 36e+6 | 0.017 | 0.524 | 0.475 | 0.175 | 0.057 | 0.003 |
| Algeria | 43e+6 | 0.017 | 0.448 | 0.332 | 0.136 | 0.051 | 0.008 |
| Tunisia | 12e+6 | 0.055 | 0.675 | 0.407 | 0.139 | 0.038 | 0.001 |
| Egypt | 100e+6 | 0.032 | 0.379 | 0.0.249 | 0.0.479 | 0.003 | 0.007 |
| South Africa | 56e+6 | 0.008 | 0.424 | 0.461 | 0.169 | 0.049 | 0.002 |
| France | 65e+6 | 0.022 | 0.799 | 0.349 | 0.173 | 0.015 | 0.009 |
| Italy | 60.48e+6 | 0.028 | 0.954 | 0.347 | 0.174 | 0.028 | 0.006 |

The estimated parameter values are strongly linked to the discipline of the local population, public health systems and the severity of the lockdown measures. Although there is so far no gold standard for assessing the accuracy of the model parameters, the values of the obtained parameters are in good agreement with the estimates given by [4, 27, 28]. The protection rate is expected to remain very close to zero since there is currently no vaccine against COVID-19 [20]. It can be seen from table 1 that Tunisia shows the highest protection rates, which reflects the increase of people infected with the new coronavirus after the opening of the country’s borders. The infection rate *β* values are in the range of the recent estimate by [4]. The lower it is, the less an infected individual will be able to infect healthy individuals. However, Tunisia shows a high infectious potential of the disease, compared to other African countries, which confirms the result of the protection rate. The obtained latent rates (Table 1) are, also, in agreement with the existing report [24], where Egypt has the longest latency period, which explains the number of infected people exceeding 94,000 cases. This may be due the delay in taking control and prevention measures against COVID-19. Notes that the latent period for COVID-19 is limited to *γ* ∈[0.2 : 0.5], so a latency period between 2 and 5 days [24]. While, Diekmann, Heesterbeek [22] notes that the average incubation period *γ*^−1^ + *δ*^−1^ for COVID-19 is between 2.1 and 11.1 days, this implies an infection rate limited to *δ* ∈[0.1:1]. Therefore, a quarantine period between 1 and 10 days. The estimation values obtained are within this estimation range, where the smallest quarantine period is recorded in Egypt. However, the parameters *λ*_*t*_ and *κ*_*t*_ governing, respectively, the recovery rate and the mortality rate of the disease, seems quite similar in the different countries, except for Egypt and Algeria. It is interesting to note that the smallest value of *λ*_*t*_ is recorded in Egypt compared to other countries, which coincides with the results of the quarantine time obtained (Table 1). This is probably due to an overload of the health system which implies a shorter average quarantine period. This reflects the poor control of virus spreading by the low testing capacity and the non-compliance with health security measures against coronavirus, especially in the most affected areas of the country. On the other hand, Algeria has the highest value of the parameter *κ*_*t*_, probably due to prevention and control measures as well as the dramatic improvement in medical conditions, such as the use new treatment protocol as well as the strict compliance with lockdown. The use of the quarantine considered by the current SEIQRDP model is based on the separation of the healthy population from the asymptomatic population in order to prevent the spread of the virus more quickly.

Figure 2 shows the effective reproduction number (*R*_*t*_) as a function of time for the different modeled countries for the same period. Analyzing the effective reproduction number (*R*_*t*_) can give us more information about how the pandemic dynamics unfolded and detecting in real time the climax or turning point. Moreover, this figure is an indication of the speed and the severity of the preventive measures as well as quarantine, adopted to reduce the average infectious period by isolating certain infectious, so that they do not transmit the infection.

**Figure 2.**
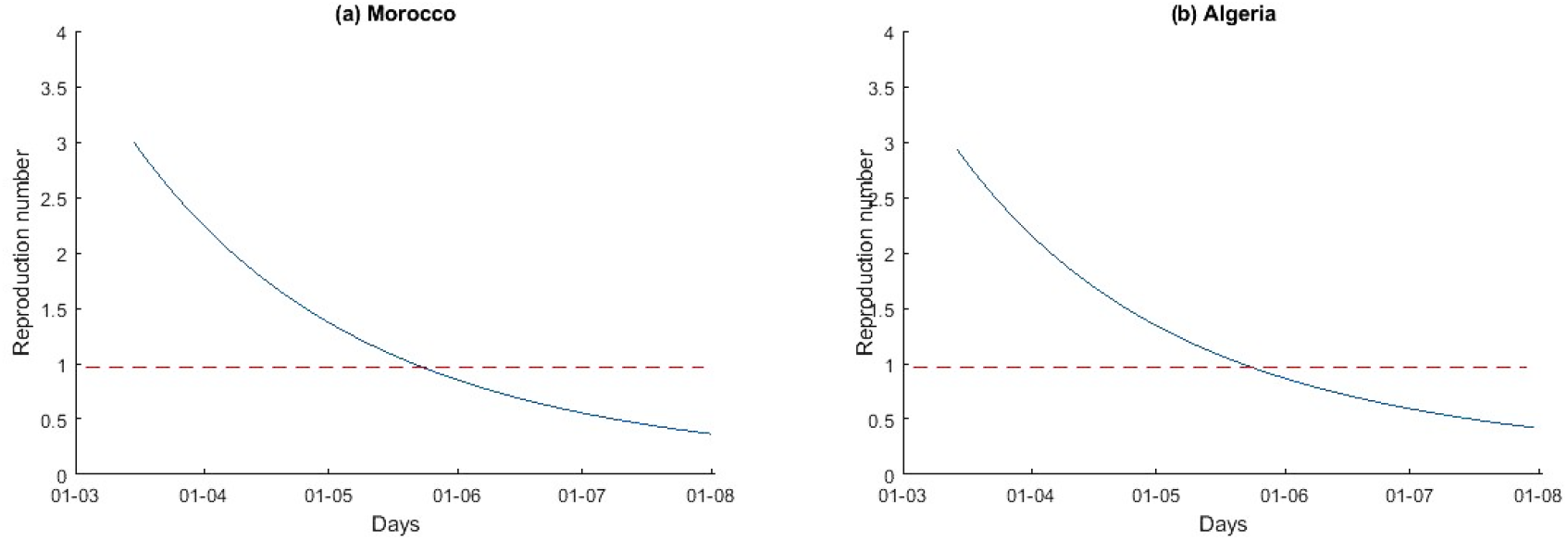
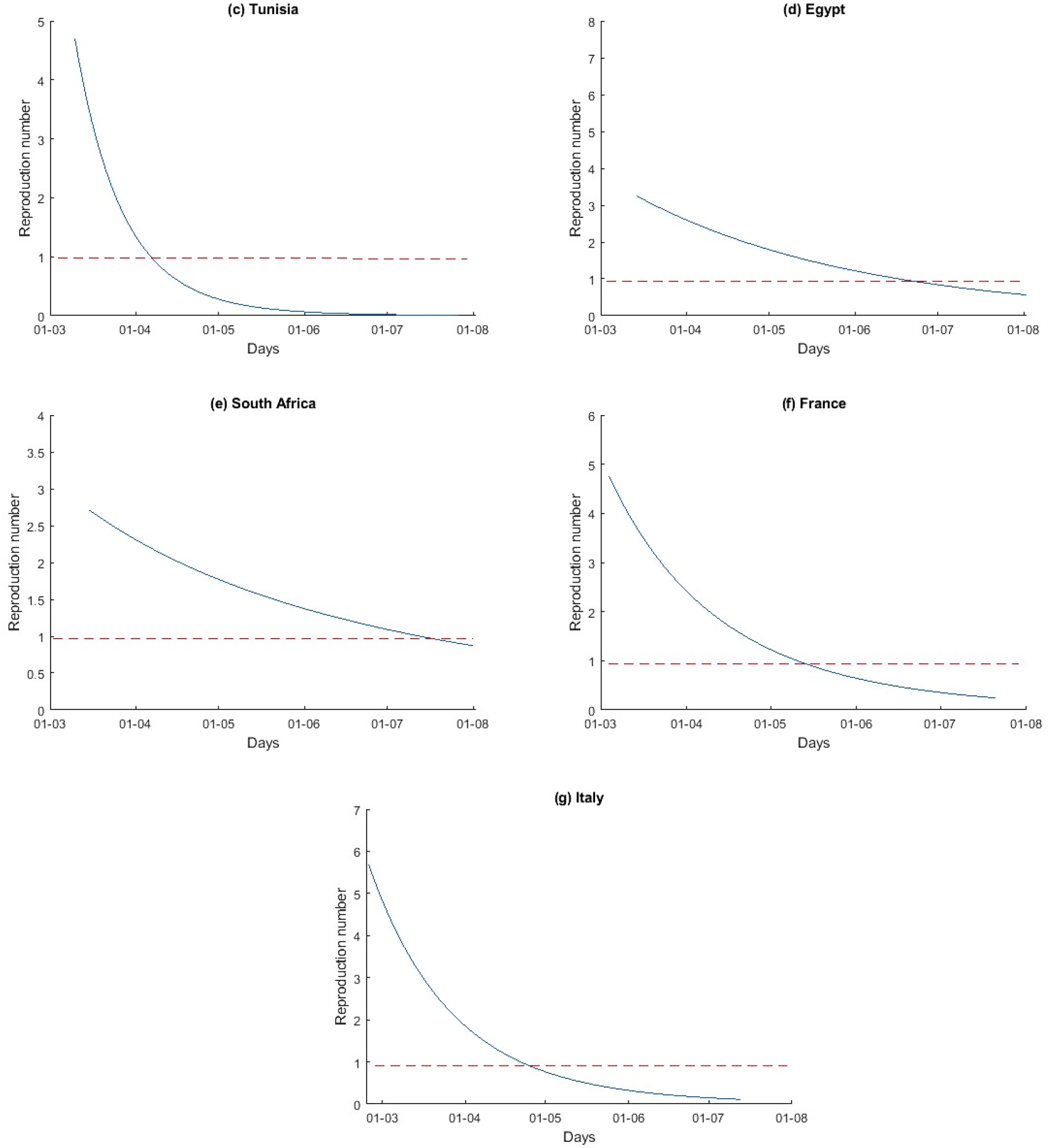
Estimated effectif reproduction number over time.

As can be seen, R_t_ becomes less than 1 between early April and early June, about four to seven weeks after the lockdown in the Maghreb countries (figure 2a-c). In fact, this shows the effects of the lockdown, the quarantine and the strict curfews adopted to reduce the effect of virus transmission, while for Egypt and South Africa, the R_t_ has not crossed the line R_t_ = 1 (mitigation phase), till end of June for Egypt and mid-July for South Africa. However, the lowest infection kinetics is recorded in Tunisia and has generated a first R_0_ = 4.690, much lower than those of other African countries and that of Italy especially. For Egypt, the number R_t_ decreased more slowly due to relative delayed lockdown compared to the other countries and an early lifting of the lockdown. However, the number of effective reproduction number depends on the parameter δ, which governs the rate of transfer from the infectious class to the quarantine class. Therefore, the use of quarantine to control the disease not only decreases the size of the endemic infectious class, but also reduces the reproduction number R_t_ below 1, so that the disease vanishes.

The estimation results (Table 1 and Figure 2) indicate that the measures taken to prevent, and control the epidemic taken by the governments have strengthened over time. Indeed, the admission and follow-up of suspect cases, quarantine and treatment of confirmed cases significantly affect the parameters values. As a result, the number of active (quarantined), suspect, cured cases and the deaths predicted by the SEIQRDP model tend to decrease over time. On the other hand, the accuracy of long-term forecasts will also decrease.

Figure 3(a-g) present the forecasts of actives (quarantined), recovery and death cases of the COVID-19 pandemic in the considered African countries and European depending on quarantine policy considered by the SEIQRDP model. While, Figure 4 presents the forecasts of the COVID-19 pandemic for France, Algeria, Morocco and Tunisia for the period after the lifting of the lockdown.

**Figure 3.**
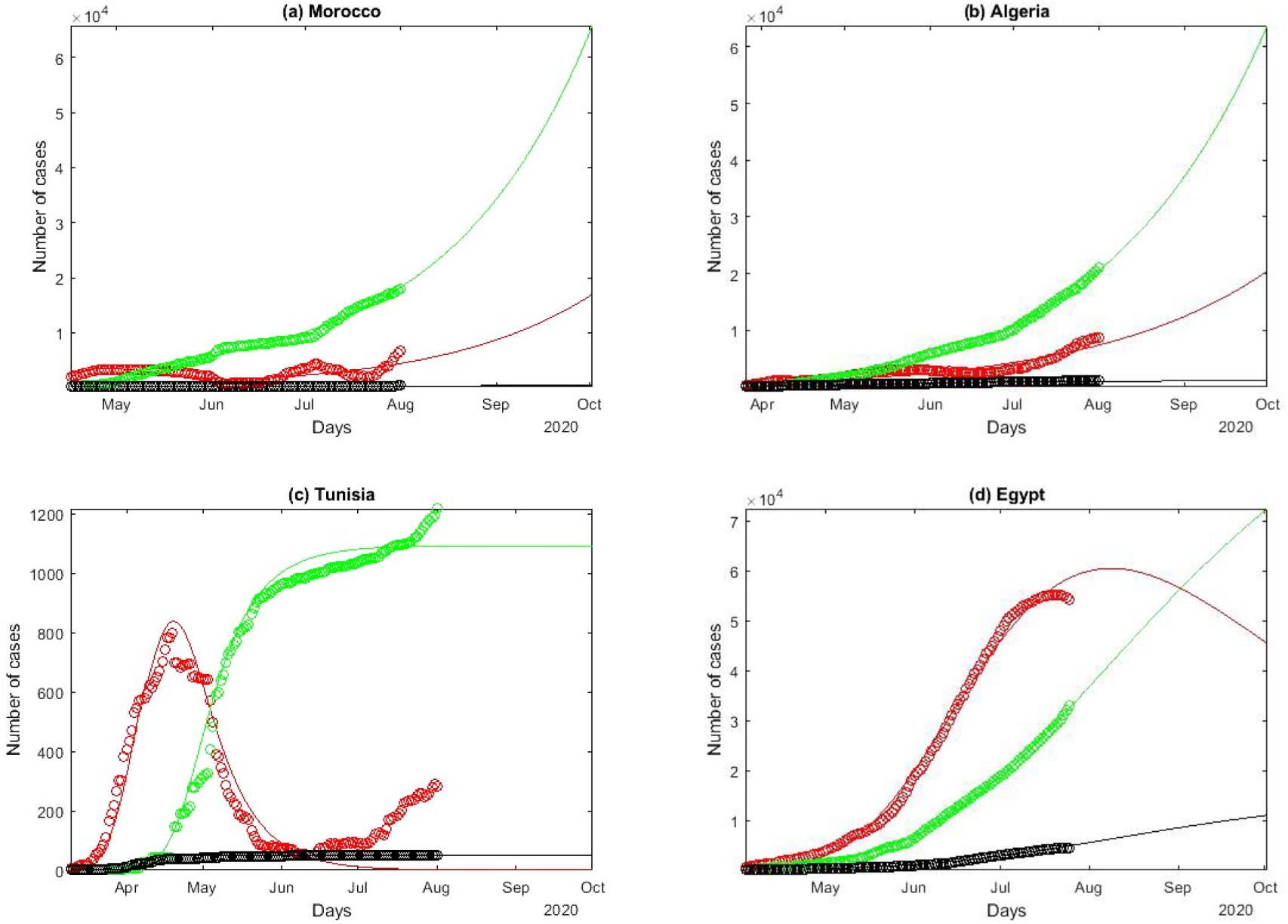
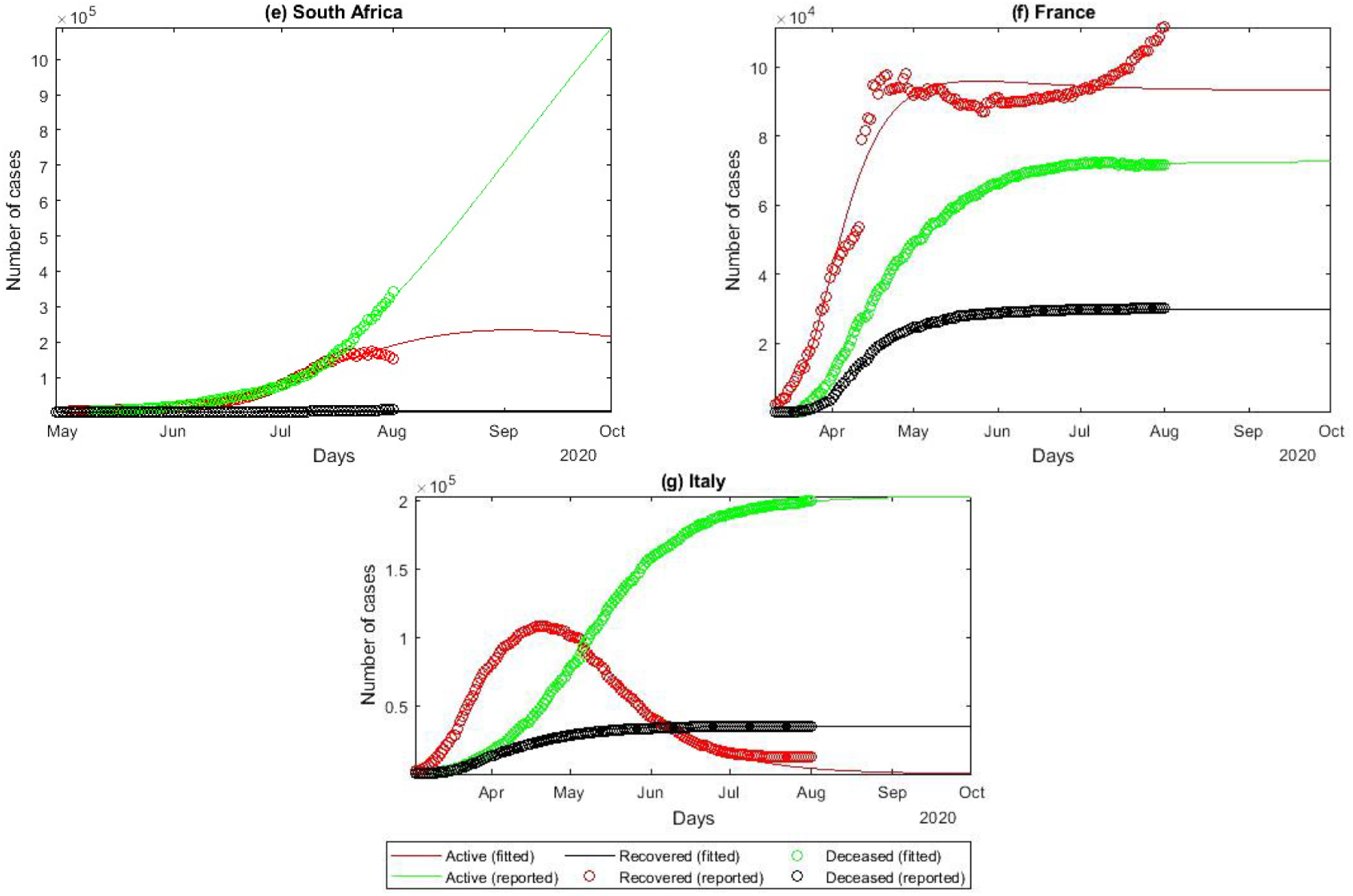
COVID-19 epidemiologic data reported and fitted by SEIRQDP model.

**Figure 4.**
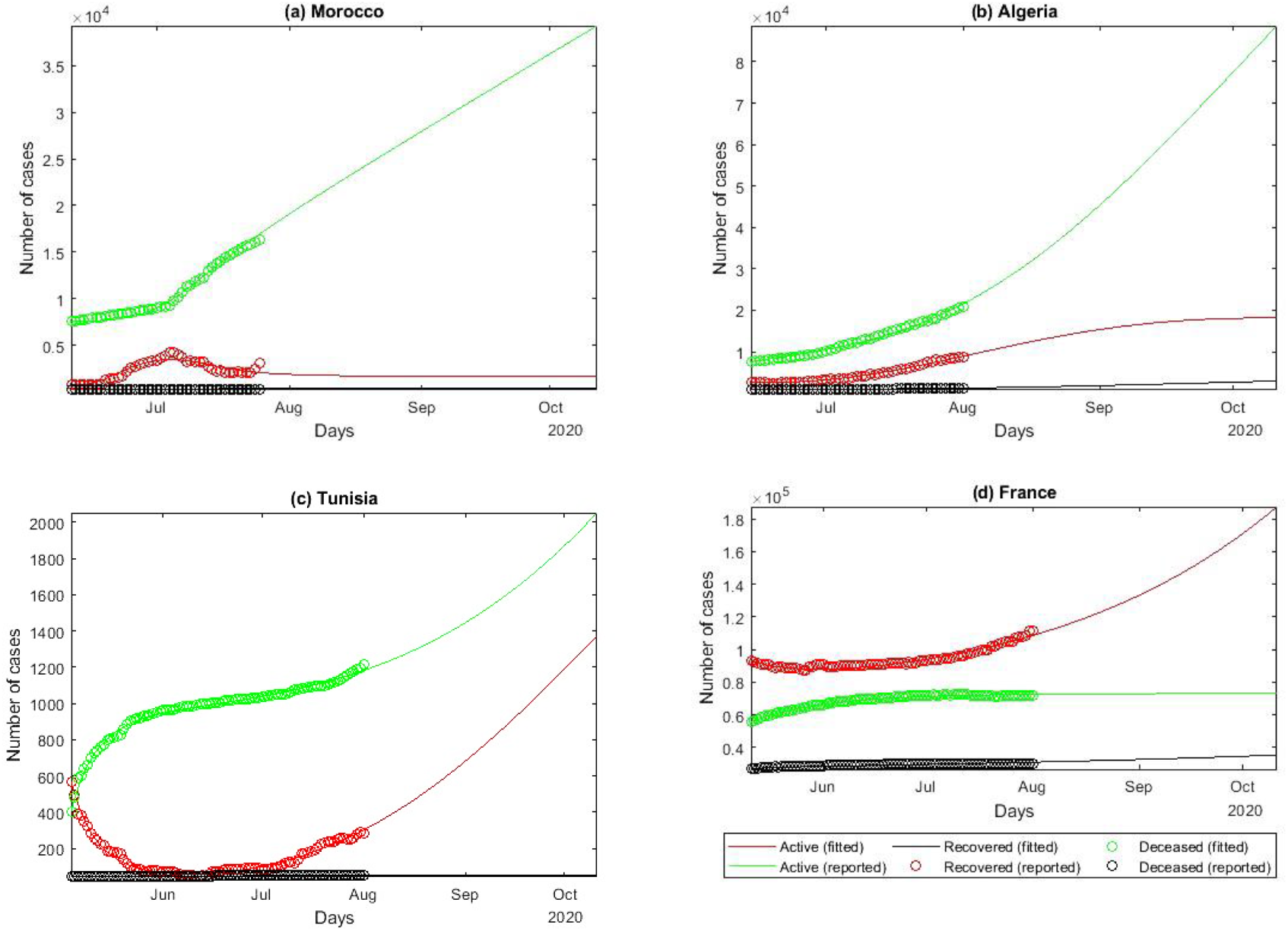
COVID-19 epidemiologic data reported and fitted by SEIRQDP model after the lockdown lifting.

Figure 3 and Figure 4 show the potential of the SEIQRDP model to predict the trend of pandemic dynamics. As can be seen from Figure 3, the curves follow roughly the same trend except for the two most affected African countries, namely, Egypt and South Africa. From Figure 4, we note that the curves follow the same trend to those of Figure 3, where we notice some non-homogeneity of the data. The curves show the potential of the model to predict the trend of pandemic dynamics. Consequently, for the forecasts of the pandemic dynamics of Algeria, Morocco, Tunisia and France, we consider the results appearing in Figure 4.

As Figure 3 shows, we find that there is a good fit between the model fitting results and the actual COVID-19 data in each country under the optimized SEIQRDP model. Therefore, it could be considered relatively reasonable to use the optimal model to predict the current epidemic situation in other countries. In addition, it is estimated that countries lockdown and closure strategies would have avoided considerable loss and millions of infections. These results show that these measures are important and necessary to respond to this serious public health emergency.

For some countries, namely, France, Algeria, Morocco and Tunisia, we notice a bad adjustment between the results of the SEIQRDP model and the actual COVID-19 data, after the lockdown lifting given by the following dates, 11 / 05, 14/06, 11/06, 04/05/2020, respectively. This can be explained by the ignorance of health security measures after the lockdown lifting. In addition, the present model, namely the SEIQRDP model, does not take into account such shocks or changes in the situation, given the very limited testing capacity, infected people can spread the virus if they do not comply with health security measures and they are not quarantined, remember that this component is very important in the model, and ignorance of the latter can affect the results of the model. Therefore, we only consider the data after the lockdown lifting to plot the forecasts of the countries in question. Figure 4 present the forecasts of active cases, recovery and death from the evolution of the COVID-19 pandemic by the SEIQRDP model adapted after the lockdown lifting.

Based on this model, the epidemic in Italy is in its final phase and should end by September. Other countries like Morocco and Algeria are expected to get rid of COVID-19 by the end of October. States like Egypt, South Africa must wait until the end of this epidemic until November. While for Tunisia and France they experience a considerable increase in the number of infected cases and an end to the epidemic is not predictable at this time. However, these forecasts would still be accurate over a short period, given the sensitivity of the model to the value of *β*, which in turn also depends on the number of contacts per person and per unit of time. This implies that the number of cases in the future could decrease or increase considerably depending on the health security measures and the respect of these measures by the population.

The Maghreb has been relatively spared by the pandemic of the new coronavirus, compared to Europe, especially, the most affected (France and Italy).

These Maghreb countries took precautionary measures very early on with border closings, general lockdown, social distancing or the wearing of protective masks. These proactive measures, dictated by the fragility of the hospital system in each country, have so far prevented the epidemic tsunami noted in France but also in Italy, as shown in Figure 3(f-g), where the peak of infected people reached around 10E6 infected in France and Italy, compared to the epidemiological situation is generally reassuring in the Maghreb nevertheless hides trajectories which diverge appreciably, after having been similar for a long time. Indeed, with more than 30.900 confirmed and 1.200 deaths cases, Algeria is the country with the most worrying situation in the Maghreb. Indeed, from Figure 3b, the number of cases confirmed daily begins to decrease after reaching its maximum. We note that there was a slight decrease in daily cases in April which seems to be explained by a low number of tests and the random rules of the lockdown. With approximately 6.500 tests performed, only cases with advanced symptoms appear to be counted. After this plateau of active cases in April, there was a worrying return until the end of May. However, according to Algerian COVID-19 forecasts by the SEIQRDP model (Figure 4b), the situation will begin to stabilize. the number of recovery cases will increase exponentially, and the number of death cases will also stabilize despite the fact that it has not grown significantly compared to the European countries. The gradually flattening in deaths is may be due to the use of the chloroquine treatment protocol [29].

However, Morocco will soon cross the threshold of 25.015 positive cases with more than 360 deaths. After a peak around early May, a decline in late May, the resumption of daily cases in early May worried the health authorities. This can be explained by the non-compliance with the rules of social distancing rules and sanitary rules during groupings, it should be noted that most new cases were attributed to contact cases. Regarding the forecast for Morocco (Figure 4a), the curves of the recovered cases seem to increase contrary to the curves of infected after the lockdown lifting. Indeed, the country has allowed the reopening of cafes, restaurants and sports halls as well as the resumption of domestic tourism and interurban travel. On the other hand, the death curve tends to zero. This encouraging result may be explained by a better knowledge of the disease [30].

With just over 1.550 cases tested positive and around 51 deaths, Tunisia is the least affected country of the three Maghreb countries by the pandemic. According to Figure 3c and Figure 4c, there is a decrease in active cases after a peak in mid-April, followed by a second significant peak predicted approximately in mid-October, due to the lockdown lifting. The curve of active cases (Figures 3c and 4c) describes well this reassuring trend, with a peak in mid-April and a drop which accelerated at the beginning of May to reach 0. This confirms the adequacy of the proposed SEIQRDP model. In addition, we notice a lower mortality rate compared to other countries as well as a significant recovery rate coupled with the low number of new cases which have eased the pressure on the hospital system. As for its Maghreb neighbors, Tunisia resorted to a treatment based on chloroquine [31]. However, according to the result of forecasts by the SEIQRDP model, the results are encouraging and confirms the end of the health crisis with a few scattered daily cases by the early August. However, for Egypt and South Africa (Figure 3d-f), the disease manifests itself in different ways compared to the Maghreb. Indeed, we note that the trend of the curves is almost the same from mid-March to July. Nevertheless, the two countries already stood out with a higher number of cases than the other countries during this same period.

was in the first place in terms of death until mid-July when the south of Africa is now the country most affected by the coronavirus in terms of death

Egypt currently has nearly 94.320 cases and around 4.830 deaths for a population of 100 million inhabitants. Until mid-July, the Egypt recorded the highest number of deaths, afterwards, South Africa became the most affected country in terms of deaths, despite the rigorous preventive measures. The situation in Egypt may be explained by the poor health system [32] and the wave of patients COVID-19 which is growing day after day. However, the results of the SEIQRDP models correspond very well to the provided data. Regarding the forecasts of the pandemic in Egypt (Figure 3d), the results show that the number of infected cases continues to increase rapidly to reach the peak around the end of August. While, the number of recovered cases will increase less rapidly, whereas, the number of deaths cases remains high compared to the Maghreb and continue to increase. This stabilization in death cases may be explained by the recent use of the drug Remdesivir to treat the patients [33]. This treatment was authorized a few days ago in the United States and Japan, based on a positive American clinical trial[34].

South Africa tops the list of countries most affected by coronavirus (in terms of contamination). To date, there have been more than 503.290 cases of COVID-19 and around 8.153 deaths. From Figure 3e, we note that infected cases have experienced significant increase since the beginning of May and continues to increase. This is due to the lockdown lifting for certain industrial sectors, with a 1.5 million people who have been authorized to return to work, under strict health protection. Whereas, before these, the situation was under control because the very strict measures taken from the beginning made it possible to slow the progression of the pandemic of coronavirus. However, the curve of restored cases continues to grow and exceeds the infected cases since mid-May despite that the latter continues to increase. This can be explained by the community screening and testing (CST) operation in working-class neighborhoods. This massive campaign made it possible to isolate infected people as quickly as possible and trace their contacts. This massive campaign partially explains South Africa’s success in the very slow development of the epidemic compared to other African countries for the same period (Figure 3a-e). Regarding the number of deaths, we note that it is stable so far, this stability is explained using the treatment protocol with chloroquine and the CST. In addition, the experience of the Ebola epidemic in South Africa which taught caregivers and populations the best practices in place, such as, isolation of the sick, precautions during care and basic hygiene such as hand washing. For the forecast of the COVID-19 pandemic in South Africa by the SEIQRDP model (Figure 3e), the situation will stabilize in terms of infected and recovered cases, while the dead experienced a certain increase in mid-July, due to the lifting of the lockdown of certain economic [35].

## 4. Conclusion

In this paper, mathematical model for the coronavirus COVID-19 epidemic development is performed for the most affected African countries based on an epidemiological SEIQRDP model, which is an adaptation of the classic SIR model widely used in mathematical epidemiology. The SEIQRDP model takes quarantine into account to match with the medical security measures adapted by many countries as an effective way of preventing the spread of the coronavirus COVID-19. Furthermore, considering all major parameters of the progression of the coronavirus, several analytic results established, while proving all the necessary properties for epidemiological relevance. This model allowed us to estimate the main epidemic parameters and forecast the evolution of the reproduction number of the disease over time in order to assess the epidemic situation and the effect of the applied preventive health measures. An acceptable agreement between the statistical data and the curves of the model was established. This indicates the suitability of the SEIQRDP in modelling the spread of the COVID-19 disease for the considered countries.

On the other hand, according to the statistical, fitted and predicted curves, we noticed that the epidemic peaks are between mid-April and mid-August, after which the epidemic declines. This situation can be explained by the policy adopted by African countries, aimed at effectively combating this virus. In addition, this confirms the effectiveness of the lockdown measures and curfews taken early enough to slow the spread of the disease. Most countries have implemented these measures as soon as the first case is detected. In France, it took 52 days after the first case to take such measures. In addition to this factor, there are many other factors that contribute to the robust and rapid response of these countries. The other main explanation is the youth of the African population, around 60% of the population under 25 years of age. In addition, the population density is lower in Africa, thus limiting the spread of the virus, as is the low mobility of African populations in comparison with Western populations, where most of the cases remain concentrated in the capitals and large cities.

However, caution should probably follow. A general lockdown lifting seems unlikely, while the gradual lifting of lockdown experienced by the countries under study by city, depending on the mortality rate and the number of new infections observed, may be reasonable in compliance with the measures of security. However, Algeria and Tunisia may experience new peaks expected mid-October, due to the lifting of lockdown, where countries have allowed the reopening of cafes, restaurants and gymnasiums as well as the resumption of tourism domestic and interurban. Nevertheless, the three Maghreb countries will have to strengthen the control of health measures, to avoid the epidemic resumption. The crucial element will remain the generalization of tests which will remain one of the main ingredients in the fight against epidemics in the country. All these precautions will effectively reduce the number of basic reproductions at each interaction, which is considered a crucial parameter during a pandemic used to estimate the risk of an outbreak of COVID-19 and assess the effectiveness of the measures implemented.

Finally, the epidemiological situation is reassuring in the three Maghreb countries, while for South Africa, the situation seems controllable, mainly due to the CST operation. However, in Egypt the number of infected cases continues to increase, the situation comes mainly from ignorance of physical distancing measures and the violation of curfews after the last relaxation of the restrictions in mid-March. In fact, without control of the population and lack of effective treatment and vaccine, there will be no way to prevent the death of many people. Furthermore, hospitals will be flooded and will never be able to manage the pandemic.

## Data Availability

COVID-19 Data Repository by the Center for Systems Science and Engineering (CSSE) at Johns Hopkins University

https://github.com/CSSEGISandData/COVID-19

## Notes

### Competing Interest Statement

The authors have declared no competing interest.

### Funding Statement

There are no funding supported in this work

